# GWAS of preeclampsia and hypertensive disorders of pregnancy uncovers genes related to cardiometabolic, endothelial and placental function

**DOI:** 10.1101/2022.05.19.22275002

**Authors:** Jaakko S. Tyrmi, Tea Kaartokallio, Inkeri Lokki, Tiina Jääskeläinen, Eija Kortelainen, Sanni Ruotsalainen, Juha Karjalainen, Samuli Ripatti, FINNPEC Study Group, FinnGen, Estonian Biobank Research Team, Triin Laisk, Johannes Kettunen, Anneli Pouta, Katja Kivinen, Eero Kajantie, Seppo Heinonen, Juha Kere, Hannele Laivuori

## Abstract

Preeclampsia is a vascular pregnancy disorder that affects 3-5% of all pregnancies. Genetic contribution to preeclampsia susceptibility is well established, but the actual risk loci have remained largely unknown. To make further discoveries of the underlying genetic architecture, we performed a new genome-wide association study (GWAS) for maternal preeclampsia and for two other combination phenotypes encompassing maternal preeclampsia and other types of gestational hypertension disorders. We combined the data resources of the Finnish pre-eclampsia cohort ‘FINNPEC’, the Finnish FinnGen project and the Estonian Biobank to obtain cases for the three abovementioned phenotypes. In addition, we performed meta-analyses of the preeclampsia phenotype combining results with the previous largest GWAS results. The controls for each phenotype comprised all parous women in the cohorts not diagnosed with these conditions. In total, we found 18 genome-wide significant associations, of which 12 have not been associated with preeclampsia in any previous maternal GWAS for maternal preeclampsia. Seven of the novel loci were near genes previously associated with blood pressure traits – supporting the concept of pregnancy as a window to future cardiovascular health. The genetic susceptibility to cardiovascular disease may manifest for the first time during pregnancy. Alterations in the integrity of the endothelium or specifically in the glomerular filtration barrier may modify disease susceptibility. Interesting novel associations are in proximity of genes involved in the development of placenta, remodeling of uterine spiral arteries and maintenance of proteostasis in pregnancy serum. Overall, the novel associated genes shed more light on the pathophysiology of preeclampsia.

## Introduction

Preeclampsia is a vascular pregnancy disorder that affects 3-5% of all pregnancies^1,2^. The disorder develops only in the presence of a placenta, and especially the early-onset preeclampsia is often accompanied by defects in the placental development and function^3,4^. Substances released by the placenta, such as antiangiogenic factors, are thought to evoke the systemic endothelial dysfunction manifested as the maternal preeclampsia symptoms including hypertension and proteinuria^5,6^. Preeclampsia is, however, both phenotypically and etiologically heterogeneous^7^. The disorder often develops without any evident placental malfunction, and the predisposition to preeclampsia is likely affected by multiple underlying cardiometabolic factors that modify a woman’s response to the pregnancy-induced stress^8^.

Genetic contribution to preeclampsia susceptibility is well established^9,10^, but the actual risk loci have remained mostly unknown. The largest genome-wide association study (GWAS) of preeclampsia by the InterPregGen consortium identified five risk loci, all of which have previously been connected to hypertension^11^. The same study also reported that the polygenic risk score for hypertension was associated with preeclampsia, implying that the genetic risk factors of these conditions are partially shared. In accordance with these observations, epidemiological evidence shows that prior cardiovascular disease inflates the risk of preeclampsia, and in addition, the previously preeclamptic women are at increased risk of developing cardiovascular disease later in life^12–15^. However, comparison with gestational hypertension in the InterPregGen study indicated that in addition to the genetic risk variants of hypertension, other variants are likely to modify susceptibility to preeclampsia^11^.

Preeclampsia is a syndromic disorder that shares features with other hypertensive disorders of pregnancy and, on the other hand, with placental disorders such as fetal growth restriction. Indeed, preeclampsia is likely to consist of several subtypes with differing etiologies^8^. The overlap in the genetic risk factors between preeclampsia and the related disorders could be subtype-specific, some subtypes sharing more features with hypertensive diseases and others being more closely linked to disorders of placental development. In this work we have therefore selected three separate phenotype groups for examination: 1) preeclampsia, 2) preeclampsia or any other type of gestational hypertension 3) preeclampsia or indication of fetal growth restriction.

The aim of our study was to identify genetic risk factors related to preeclampsia and hypertensive disorders of pregnancy in the maternal genome-wide meta-analysis comprising samples from the closely related populations of Finland and Estonia. For the strict pre-eclampsia, these data were supplemented with the previously published InterPregGen GWAS to achieve the largest genetic data set studied in preeclampsia to date. In addition, we performed GWAS in smaller paternal and fetal sample sets from Finland to identify risk loci of preeclampsia conveyed via the fetus. Our functional annotation of the associated loci suggested putative variants and genes likely contributing to the pathophysiology of preeclampsia and other hypertensive disorders of pregnancy.

## Material & Methods

### Ethical considerations

The FINNPEC study was approved by the Coordinating Ethics Committee of the Hospital District of Helsinki and Uusimaa (149/EO/2007). All FINNPEC study participants and the participating parents of the neonates provided a written informed consent.

Patients and control subjects in FinnGen provided informed consent for biobank research, based on the Finnish Biobank Act. Alternatively, older research cohorts, collected prior the start of FinnGen (in August 2017), were collected based on study-specific consents and later transferred to the Finnish biobanks after approval by Fimea, the National Supervisory Authority for Welfare and Health. Recruitment protocols followed the biobank protocols approved by Fimea. The Coordinating Ethics Committee of the Hospital District of Helsinki and Uusimaa (HUS) approved the FinnGen study protocol Nr HUS/990/2017.

The FinnGen study is approved by the Finnish Institute for Health and Welfare (permit numbers: THL/2031/6.02.00/2017, THL/1101/5.05.00/2017, THL/341/6.02.00/2018, THL/2222/6.02.00/2018, THL/283/6.02.00/2019, THL/1721/5.05.00/2019, THL/1524/5.05.00/2020, and THL/2364/14.02/2020), the Digital and population data service agency (permit numbers: VRK43431/2017-3, VRK/6909/2018-3, VRK/4415/2019-3), the Social Insurance Institution (permit numbers: KELA 58/522/2017, KELA 131/522/2018, KELA 70/522/2019, KELA 98/522/2019, KELA 138/522/2019, KELA 2/522/2020, KELA 16/522/2020 and the Statistics Finland (permit numbers: TK-53-1041-17 and TK-53-90-20).

The Biobank Access Decisions for the FinnGen samples and the data utilized in the FinnGen Data Freeze 6 include: THL Biobank BB2017_55, BB2017_111, BB2018_19, BB_2018_34, BB_2018_67, BB2018_71, BB2019_7, BB2019_8, BB2019_26, BB2020_1, Finnish Red Cross Blood Service Biobank 7.12.2017, Helsinki Biobank HUS/359/2017, Auria Biobank AB17-5154, Biobank Borealis of Northern Finland_2017_1013, Biobank of Eastern Finland 1186/2018, Finnish Clinical Biobank Tampere MH0004, Central Finland Biobank 1-2017, and Terveystalo Biobank STB 2018001.

The Estonian Biobank (EstBB) is a population-based biobank that has obtained clinical data from the national registries and hospital databases. Analyses in the EstBB were carried out under ethical approval 1.1-12/624 from the Estonian Committee on Bioethics, and Human Research and data release N05 from the EstBB. All biobank participants have signed a broad informed consent form.

### Study phenotypes and cohorts

For the maternal meta-analysis, genome-wide genotyped and imputed samples from the FINNPEC, FinnGen and EstBB were utilised. In addition, we included summary statistics from the earlier largest meta-analyses study of preeclampsia^11^. For the fetal and paternal genome-wide association analysis, samples from the FINNPEC cohort were available. The analyses were performed with 3 phenotypes: 1) preeclampsia, eclampsia or preeclampsia superimposed on chronic hypertension (PE); 2) the aforementioned preeclampsia phenotypes, gestational hypertension or other maternal hypertension during pregnancy (PE-HTP); and 3) preeclampsia or fetal growth restriction (as mother’s diagnosis), used as proxy for small for gestational age (FinnGen and EstBB) or diagnoses of placental insufficiency or small-for-gestational age infant (FINNPEC) (PE-FGR). The case group consisted of women fulfilling these inclusion criteria while the control group for each phenotype comprised all parous women not diagnosed with these conditions. The sample sizes available in each analysis are presented in Table 1 and the study cohort and phenotypes are described in more detail below.

**Table 1.** Sample sizes in the association analyses in the FINNPEC, FinnGen, Estonian Biobank and InterPregGen studies and in the maternal meta-analyses for three phenotypes.

| **Cohort** | **Analysis** | **Case** | **Ctrl** | **Total n** |
| --- | --- | --- | --- | --- |
| FINNPEC | PE, maternal | 1479 | 972 | 2451 |
| EstBB | PE, maternal | 1464 | 38 105 | 39 569 |
| FinnGen | PE, maternal | 4285 | 83 285 | 87 570 |
| InterPregGen | PE, maternal | 9515 | 157 719 | 167 234 |
| **Meta-analysis** | **PE, maternal** | **16 743** | **280 081** | **296 824** |
| FINNPEC | HTP, maternal | 1689 | 778 | 2467 |
| EstBB | HTP, maternal | 4084 | 35 628 | 39 712 |
| FinnGen | HTP, maternal | 9427 | 78 601 | 88 028 |
| **Meta-analysis** | **HTP, maternal** | **15 200** | **115 007** | **130 207** |
| FINNPEC | PE-FGR, maternal | 1564 | 890 | 2454 |
| EstBB | PE-FGR, maternal | 2772 | 36 797 | 39 569 |
| FinnGen | PE-FGR, maternal | 6464 | 81 538 | 88 002 |
| **Meta-analysis** | **PE-FGR, maternal** | **10 800** | **119 225** | **130 025** |
| FINNPEC | PE, fetal | 796 | 894 | 1690 |
| FINNPEC | HTP, fetal | 946 | 750 | 1696 |
| FINNPEC | PE-FGR, fetal | 839 | 852 | 1691 |
| FINNPEC | PE, paternal | 595 | 654 | 1249 |
| FINNPEC | HTP, paternal | 697 | 557 | 1254 |
| FINNPEC | PE-FGR, paternal | 633 | 618 | 1251 |

PE=preeclampsia; PE-HTP=preeclampsia, gestational hypertension or preeclampsia superimposed on chronic hypertension; PE-FGR=preeclampsia or indication of fetal growth restriction.

The FINNPEC study is a nationwide preeclampsia case-control cohort recruited from five university hospitals in Finland. The cohort consist of both prospectively and retrospectively recruited women, as well as spouses and neonates of the participating women in the prospective arm of the cohort. Blood samples for DNA extraction were collected from all the participants, and first and third trimester serum samples as well as placental samples from a subset of the women. In addition, extensive clinical information collected from hospital records of the participants’ obstetric histories, pregnancy complications and outcomes, and laboratory, blood pressure and proteinuria measurements during pregnancy, as well as information on delivery and the newborn have been obtained. In the FINNPEC cohort, preeclampsia was defined according to the ACOG 2002 criteria as hypertension (≥140/90 mmHg) and proteinuria (≥0.3 g/24h, or 0.3 g/L, or two ≥1+ dipstick readings) occurring after 20 weeks of gestation. Gestational hypertension was defined as hypertension (≥140/90 mmHg) occurring after 20 weeks of gestation in the absence of proteinuria. Birth weights below −2.0 SD units according to the Finnish standards^16^ were classified as small-for-gestational age. Placental insufficiency was defined as umbilical artery resistance index or pulsatility index ≥+2 SD. Women with multiple or ovum donation pregnancy, age below 18 years, or an inability to provide informed consent based on information in Finnish or Swedish were excluded. All unaffected women not fulfilling the exclusion criteria were included in the control group. Diagnoses were established based on medical records and confirmed independently by a research nurse and a study physician. A total of 1479, 1689 and 1564 FINNPEC cases of PE, HTP and PE-FGR, respectively, were available for the maternal meta-analyses. A full list of FinnGen contributors can be found in Supplementary Data 1.

The FinnGen project combines genotype data from Finnish nationwide biobanks that are linked with digital health records from the national hospital discharge (from 1968 onwards), cancer (1953-), death (1969-) and medication reimbursement (1995-) registers. The Data Freeze 6 used in this study contains the genomic and health record data for 6% of adult Finnish women. In FinnGen, 88028 women with offspring were included in the study. The cases of each phenotype were defined according to the ICD-entries described in Table S1, with total of 4743, 9427 and 6464 cases in the PE, PE-HTP and PE-FGR phenotypes respectively. A full list of FinnGen contributors can be found in Supplementary Data 2.

EstBB is a population-based biobank, which closely reflects the age, sex and geographical distribution of the Estonian population. ICD-code information is obtained from the Estonian Causes of Death, Estonian Cancer, Estonian Tuberculosis and Estonian Health Insurance Fund registries, and also from the databases of Tartu University Hospital and North Estonia Medical Centre. The case definition for the EstBB 200K data freeze was similar to FinnGen, but only the ICD-10 codes were used. Total case counts for the PE, PE-HTP and PE-FGR phenotypes were 1464, 4084 and 2772, respectively.

In addition, we used summary statistics from the previous largest genome-wide association meta-analysis for preeclampsia conducted by Steinthorsdottir et al^11^. This dataset was generated based on 9515 preeclamptic women and 157,719 controls from Europe and Central Asia. These data were included only for the meta-analysis of the PE phenotype.

### Genotyping and imputation

FINNPEC: Genotyping of the FINNPEC samples was performed using Infinium Global Screening Array-24 v2.0 BeadChip (Illumina Inc., San Diego, CA, USA). Pre-imputation quality control was carried out with Plink 1.07 and 1.9. Duplicated samples and members of triads and dyads that did not show expected genetic relationships based on Mendelian errors and the IBD analysis with the Plink’s –genome option were excluded. In addition, samples with unresolved sex mismatch, missingness rate over 5%, heterozygoty rate ±4 SD or non-Finnish ancestry based on MDS analysis were excluded. In the variant-wise quality control, variants with missing call rate >2%, Hardy-Weinber equilibrium (HWE) p<1×10^−6^, or minor allele count <3 were removed. The genotyped samples were pre-phased with Eagle v2.3.5 and imputed with Beagle v4.1 using a population-specific reference panel SISu v3, which consists of 3775 whole genome sequenced individuals of Finnish ancestry.

FinnGen: Sample genotyping in FinnGen was performed using Illumina and Affymetrix arrays (Illumina Inc., San Diego, and Thermo Fisher Scientific, Santa Clara, CA, USA). Genotype calls were made using GenCall or zCall for Illumina and AxiomGT1 algorithm for Affymetrix data. Genotypes with HWE p-value p<1×10^−6^, minor allele count <3 and genotyping success rate <98 % were removed. Samples with ambiguous gender, high genotype missingness > 5% and those that were outliers in population structure (> 4 SD from mean on first two dimensions) were omitted. Samples were pre-phased with Eagle 2.3.5 using 20,000 conditioning haplotypes. Genotypes were imputed with Beagle 4.1 using SiSu v3 imputation reference panel.

Estonian Biobank: All EstBB participants were genotyped at the Core Genotyping Lab of the Institute of Genomics, University of Tartu, using Illumina GSAv1.0, GSAv2.0, and GSAv2.0_EST arrays. Samples were genotyped and PLINK format files were created using Illumina GenomeStudio v2.0.4. Individuals were excluded from the analysis if their call-rate was < 95% or if sex defined based on heterozygosity of X chromosome did not match sex in phenotype data. Before imputation, variants were filtered by call-rate < 95%, HWE p-value < 1e-4 (autosomal variants only), and minor allele frequency < 1%. All variants were changed to be from TOP strand using GSAMD-24v1-0_20011747_A1-b37.strand.RefAlt.zip files from https://www.well.ox.ac.uk/~wrayner/strand/webpage. Prephasing was done using the Eagle v2.3 software (number of conditioning haplotypes Eagle2 uses when phasing each sample was set to: --Kpbwt=20000) and imputation was done using Beagle v.28Sep18.793 with effective population size ne=20,000. Population specific imputation reference of 2297 WGS samples was used^17^.

### Association analyses

FINNPEC: The association analyses were performed with SAIGE version 0.39. The null models were adjusted for maternal age at birth and the first 10 principal components (PC).

FinnGen: Association analysis was performed using generalized mixed model as implemented in SAIGE v.0.39.1^18^. Included adjustments were age, genotyping batches and the first ten PCs.

Estonian Biobank: Association analysis was carried out using SAIGE (v0.38) software implementing mixed logistic regression model with year of birth and 10 PCs as covariates.

### Meta-analyses

For meta-analysis, the GWAS summary statistics of FinnGen were lifted over from hg38 to hg37 reference genome build using UCSC liftOver^19^ and only high imputation quality markers were retained (INFO score > 0.7). METAL software^20^ was used to perform inverse variance-weighted meta-analysis with genomic control correction applied. Genomic inflation factor estimates were calculated with ‘LD Score regression’ software^21^. Genome-wide significance was set to p < 5 × 10^−8^. The meta-analysis was conducted by two analysts independently and summary statistics were compared for consistency. Formatting and preparation of the association summary statistics data for downstream analysis was managed with the workflow management software STAPLER^22^.

### Annotation of loci

We defined annotated loci as the lead variants and the surrounding +-1 Mbp area. To identify putative candidate genes we first used the web-based FUMA platform^23^. ANNOVAR^24^ and MAGMA^25^ are integrated as part of FUMA, and were used for performing gene-based analysis and creating functional annotation for the meta-analysis results, respectively. We examined whether the variants residing in the associating loci had any predicted functional consequences by identifying missense mutations or pathogenic variants. Furthermore, we assessed if the variants had high CADD score^26^, and examined whether the variants were eQTLs based on the information provided in the Genotype-Tissue Expression (GTEx) Portal or had been previously associated with any preeclampsia related phenotype in the GWAS catalog^27^.

If the above functional annotation did not provide support for any causal variants or genes within a locus, we then conducted a broad literature search to identify additional important information of all other genes within ±1Mbp of each lead variant. Here, genes most proximal to the lead variant in each locus were prioritized. In addition to the literature search, we explored the information provided in GenBank^28^ and UniProt^29^ databases.

Lastly, we used LDSC software^21,30^ to test for potential inflation in the test statistics, estimate regression-derived SNP-based heritability and to evaluate genetic correlation (*r*_*g*_) between PE, PE-HTP and PE-FGR and 895 other previously published phenotypes (Supplemetary file 3). The phenotypes were selected to closely match the (currently defunct) LDhub^31^ dataset, combined with additional circulating metabolite data from Kettunen et al.^32^.

### Data availability

Meta-analysis summary statistics will be made available upon publication.

## Results

### Association signals from the maternal meta-analyses

Considering the syndromic nature of preeclampsia and its close resemblance to other pregnancy disorders, we conducted maternal meta-analyses with three overlapping preeclampsia phenotypes (Table 1): 1) preeclampsia (PE); 2) preeclampsia or other hypertensive pregnancy (PE-HTP); and 3) preeclampsia or fetal growth restriction (as mother’s diagnosis), used as proxy for small for gestational age (PE-FGR). In the PE, PE-HTP and PE-FGR phenotypes, we identified altogether 9, 13 and 2 genome-wide significant loci, respectively (Table 2, Figures 2 and S1-5). Four of the PE-META loci, nine of the PE-HTP-META and one of the PE-FGR-META loci had not been significantly associated with preeclampsia in the earlier maternal GWAS.

**Table 2.**
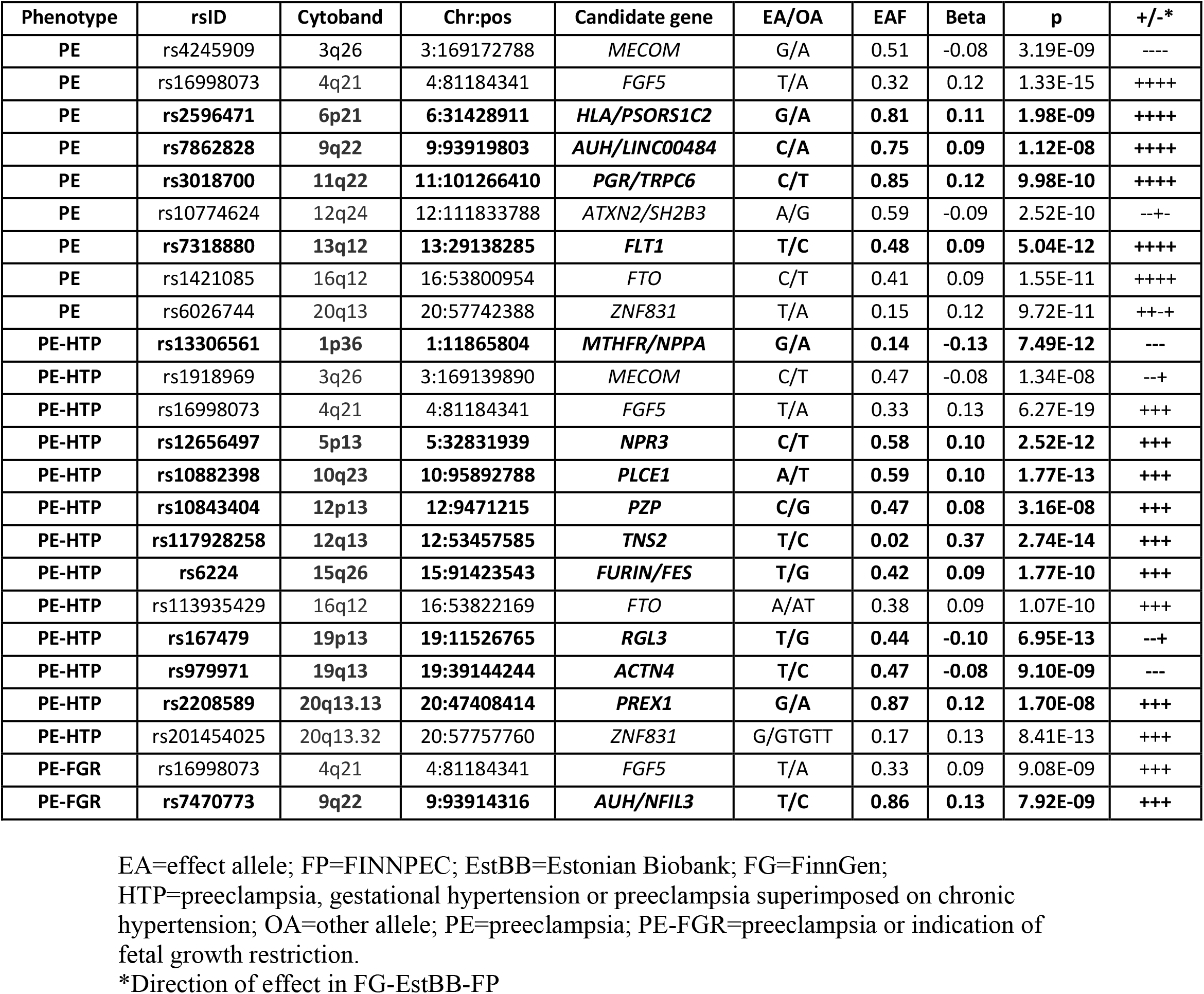
Lead variants of the genome-wide significant loci from the maternal meta-analyses in the PE, HTP and PE-FGR phenotypes. Novel loci are bolded.

**Figure 1.**
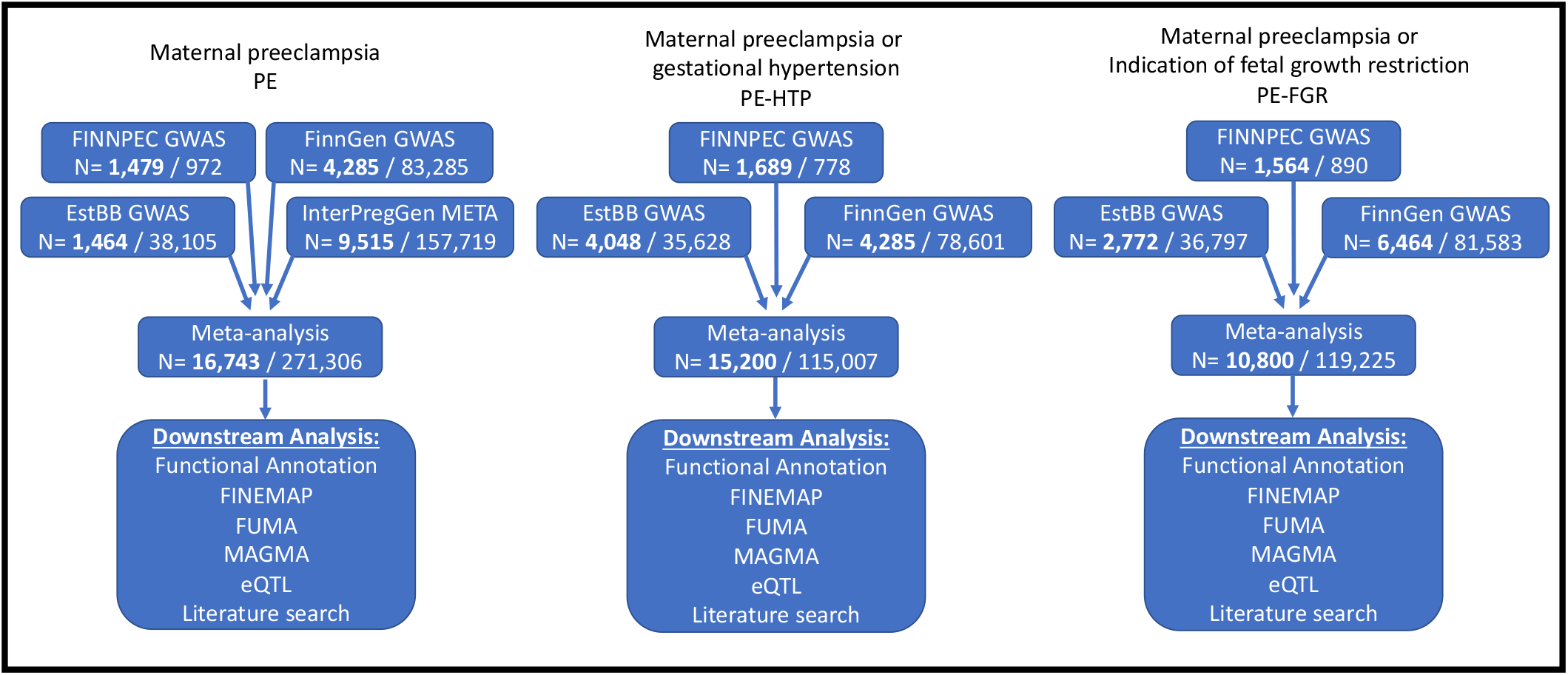
Flow chart of the study design.

**Figure 2.**
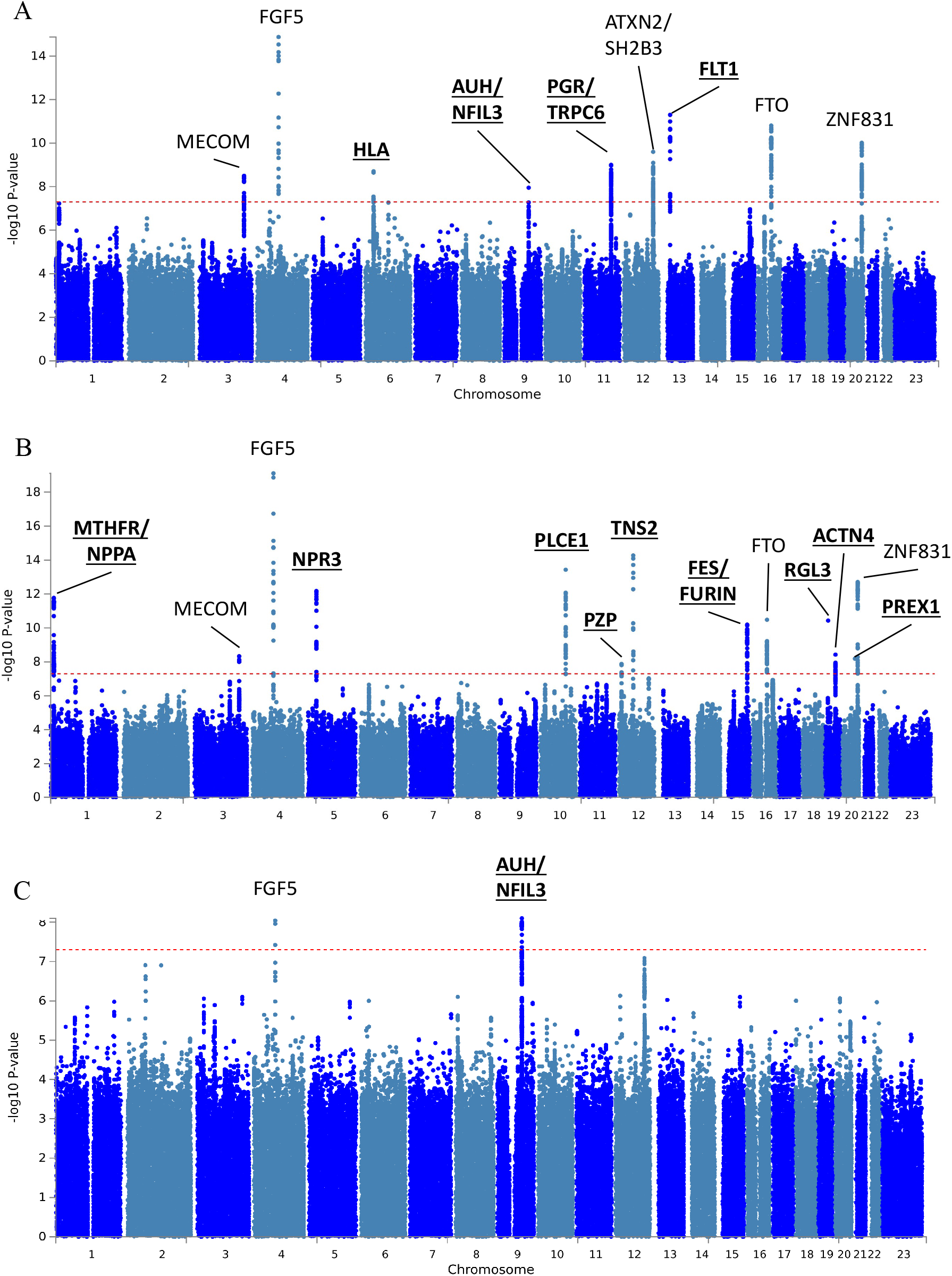
Manhattan plot for the meta-analysis results of PE (A), PE-HTP (B) and PE-FGR (C) phenotypes with genome-wide significant loci labeled with most likely candidate gene. Novel loci not detected in previous GWAS are bolded and underlined. PE=preeclampsia; PE-HTP=preeclampsia, gestational hypertension or preeclampsia superimposed on chronic hypertension; PE-FGR=preeclampsia or indication of fetal growth restriction.

There was no evidence of inflation in the meta-analysis test statistics based on calculated LD Score intercepts, which were 0.9928 (SE 0.0085), 0.9717 (SE 0.0082) and 0.9722 (SE 0.0082) for the PE, PE-HTP and PE-FGR phenotypes, respectively. The majority of the directions of the effects were concordant between the cohorts with the exception of four loci near 3q26, 12q24, 19p13, 20q13 that showed discordant effect of direction in the smallest cohort of FINNPEC to the others (Table 2). Also, the effect sizes of the lead variants were in general modest and similar across the loci, with an average beta of 0.11. We did not observe genetic heterogeneity, except for the lead variant rs167479 at locus 19p13 (heterogeneity p-value 0.004). Almost all the lead variants were common, with the only exception being the lead variant of the 12q13 locus (minor allele frequency 0.02).

### PE-META reveals several novel risk loci for preeclampsia and replicates earlier findings

Four of the nine genome wide significant loci in PE-META have not been previously identified in the maternal GWAS of preeclampsia (Figure 2A). For the first of these, *FLT1*, the earlier InterPregGen GWAS found an association between the locus in the fetal genome and mother’s preeclampsia^33^. The lead variant in the *FLT1* locus (rs7318880) is in tight linkage disequilibrium (*r*^*2*=^0.98) with the lead variant rs4769612 in Steinthorsdottir et al. ^11^. The soluble form of the *FLT1* encoded protein, sFlt1, is a well-known biomarker for preeclampsia: levels of this antiangiogenic protein are increased in the circulation of women destined to develop the disease already before the onset of the symptoms and correlate with the disease severity^6,34^. By sequencing a large number of candidate genes, we have previously found low-frequency variants in the *FLT1* gene to be protective of maternal preeclampsia^35^. The current study is the first to find a genome-wide significant association of the locus in preeclamptic mothers.

We also found a genome-wide significant association within the HLA region, which is known for its high density of genes involved in immune regulation and recognition. Tissue compatibility, autoimmunity, and cardiovascular diseases are processes with known susceptibility loci within the HLA and pathophysiological relevance in preeclampsia. In pregnancy, HLA-G, -F, -E and -C are potentially directly involved in tolerance induction by recognition of fetal trophoblast cells by maternal immune cells, the classical HLA genes likely also play a role in the maintenance of peripheral tolerance^36–38^. The association lies within the major susceptibility locus for psoriasis (PSORS1), known for accounting much of the genetic risk for psoriasis^39,40^. The two top outliers of the HLA-region in MAGMA test, ‘Coiled-Coil Alpha-Helical Rod Protein 1Coiled-Coil Alpha-Helical Rod Protein 1’ (*CCHCR1*) and ‘Psoriasis Susceptibility 1 Candidate 2’ (*PSORS1C2*), have indeed been previously suggested as risk genes for psoriasis^41–43^. Additionally, evidence shows *PSORS1C2* effects on apolipoprotein B and blood protein levels^44,45^. Psoriasis increases the risk of adverse pregnancy outcomes and specifically preeclampsia with an odds ratio of 1.5^45–48^. Furthermore, intronic regions within HLA class I were associated with preeclampsia. Other putative causal genes suggested by MAGMA analysis include *MCCD1*, previously suggested to cause cardiovascular disease^49^, *TCF19*, possibly harboring variants for type 2 diabetes^50^, and *BAG6*, which contributes to numerous cellular processes, including immunological pathways^51^. Due to the high gene density, extreme polymorphism and complex haplotype structure, identifying exact candidate genes in the HLA region typically involves specialized methods. Identifying genome-wide significant association within the HLA region in preeclamspsia is of great interest.

Another novel association was detected near the genes ‘AU RNA binding methylglutaconyl-CoA hydratase’ (*AUH*) and ‘Nuclear Factor, Interleukin 3 Regulated’ (*NFIL3*). According to the GTExPortal the lead variant rs7862828 is an eQTL for the RNA-gene *LINC00484* in multiple tissues. The gene is poorly characterized, but variants within the gene have been associated with several traits, such as waist-to-hip ratio adjusted for BMI^52^, total bilirubin levels^53^ and glycosuria in pregnancy^54^.

In another novel associating locus at 11q22, the intergenic variant rs3018700 lies near ‘progesterone receptor’ gene (*PGR*) and ‘Transient Receptor Potential Cation Channel Subfamily C Member 6’ (*TRPC6). PGR* mediates the effects of progesterone, an essential hormone in pregnancy that maintains pregnancy and promotes endometrial maturation, angiogenesis, vasodilation and placentation^55^. Transcriptomic profiling has revealed perturbed PGR signalling in the decidual endometrium of previously preeclamptic women^56^. The eQTL data provide some support for the causativity of the *PGR* gene. Mutations in the other plausible candidate gene in the same locus, *TRPC6*, contributes to various biochemical pathways throughout the body. Variants in this gene have been shown to cause familial focal segmental glomerulosclerosis^57^, and to affect vascular smooth muscle contractility^58^, which may subsequently contribute to the risk of cardiac hypertrophy, heart failure^59^ and idiopathic pulmonary arterial hypertension^60^. Also, the gene exhibits enhanced placental expression, and the *TRPC6* knock-out mice presents with structural changes of the placenta and reduced litter sizes^61^.

In addition to these novel findings, we replicated all five loci reported by Steinthorsdottir et al.^11^. The lead variant in the *FGF5* locus (rs16998073) reached genome-wide significance in all phenotypes studied here (Table 2, Figure 2), and is in high linkage disequilibrium (LD, r^2^ = 0.93) with the lead variant of the Steinthorsdottir study (rs1458038). The other replicated loci include ‘MDS1 And EVI1 Complex Locus’ (*MECOM*), ‘SH2B adaptor protein 3’ / ‘ataxin 2’ (*SH2B3* / *ATXN2*), ‘FTO Alpha-Ketoglutarate Dependent Dioxygenase’ (*FTO*) and ‘Zinc Finger Protein 831’ (*ZNF831*).

In addition, for the PE phenotype significant p-values were obtained in the MAGMA gene-based analysis for the ‘Methylenetetrahydrofolate Reductase’ (*MTHFR*) and ‘Chloride Voltage-Gated Channel 6’ (*CLCN6)* genes (Fig. 3). Variants in both of these genes have previously been associated with blood pressure in UKBB^62^. Variants in this same locus reached genome-wide significance in the meta-analysis of the PE-HTP phenotype, with the PE-HTP increasing allele also increasing the risk for blood pressure according to the UKBB data. Genetic correlations calculated with LDSC between PE and 895 phenotypes show that the most strongly associated phenotypes are related to blood pressure or various cardiovascular disease, but also to several measures of body fat (Supplementary File 3).

**Figure 3.**
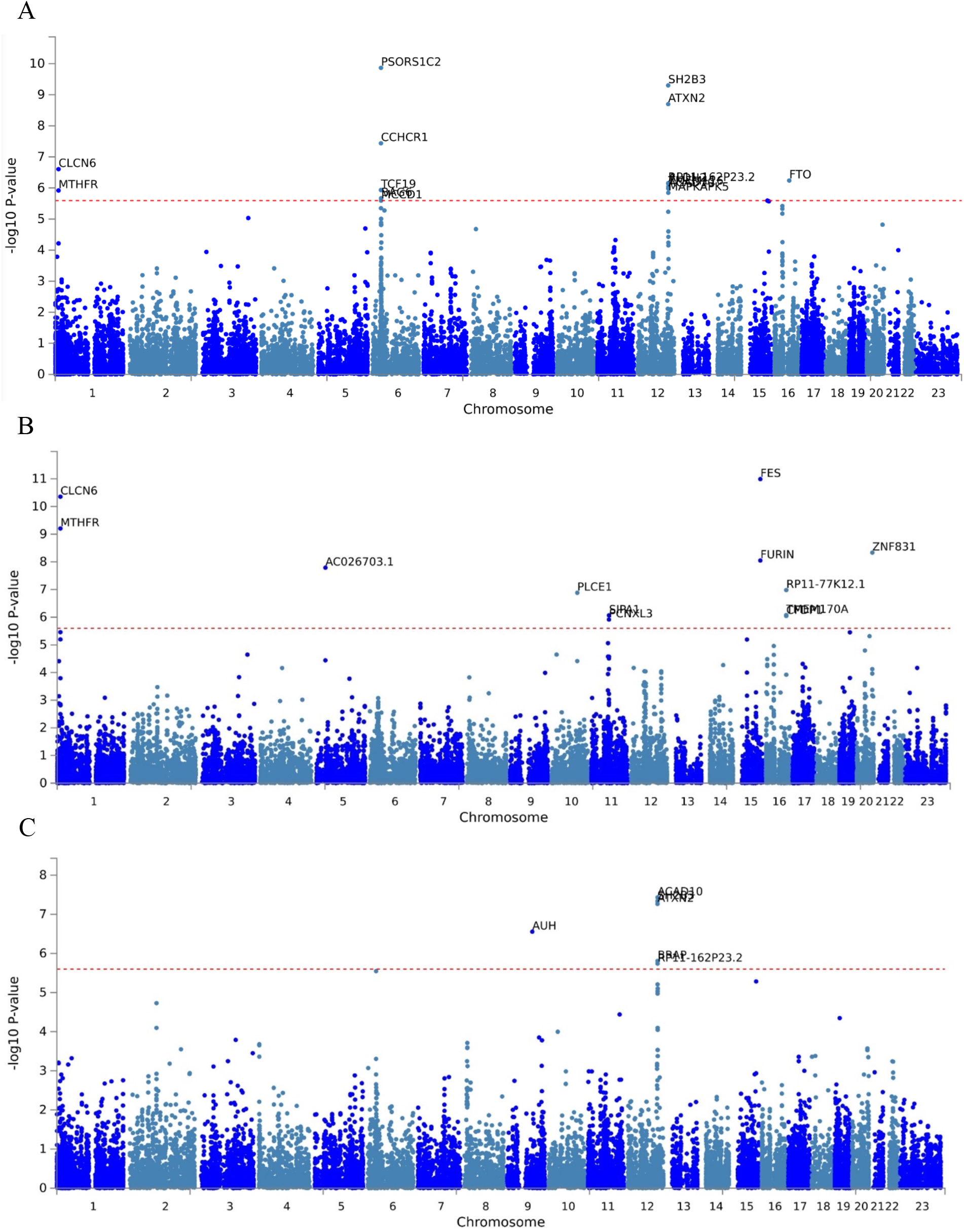
MAGMA gene based test results for the meta-analysis results PE (A), PE-HTP (B) and PE-FGR (C) phenotypes. PE=preeclampsia; PE-HTP=preeclampsia, gestational hypertension or preeclampsia superimposed on chronic hypertension; PE-FGR=preeclampsia or indication of fetal growth restriction.

### PE-HTP-META identifies novel loci with candidate genes related to hypertensive diseases and maintenance of successful pregnancy

PE-HTP-META revealed nine novel loci not identified by Steinthorsdottir or in the PE phenotype studied here. Of the identified novel associating loci, seven harbor genes previously associated with high blood pressure or hypertensive diseases. These include the *MTHFR, CLCN6* and ‘Natriuretic Peptide A’ (*NPPA*) genes on chromosome 1p36, ‘Natriuretic Peptide Receptor 3’ (*NPR3*) on 5p13, Phospholipase C Epsilon 1 (*PLCE1*) on 10q23, Tensin 2 (*TNS2*) on 12q13, ‘Furin, Paired Basic Amino Acid Cleaving Enzyme’ (*FURIN*) on 15q26, ‘Ral Guanine Nucleotide Dissociation Stimulator Like 3’ (*RGL3*) and Phosphatidylinositol-3,4,5-Trisphosphate Dependent Rac Exchange Factor 1 (*PREX1*) on 20q13. However, two loci on 12p13 and 19q13 do not appear to contain any genome-wide significant variants directly contributing to hypertensive diseases, or genes associated with hypertensive disorders in close proximity. The most likely candidate genes in these two loci are ‘pregnancy zone protein’ (*PZP*) on 12p13 and ‘actinin alpha 4’ (*ACTN4*) on 19q13.

*PZP* is a protease inhibitor that prevents the activity of all four classes of proteases and stabilises misfolded proteins, which have been shown to accumulate in preeclampsia and contribute to its pathophysiology^63–67^. For instance, *PZP* has been shown to inhibit the aggregation of amyloid beta peptide^68^ – an important driver in plaque formation in several disorders, including preeclampsia, age-related macular degeneration (particularly in women), and Alzheimer disease. Also importantly, the protein has been suggested to clear out pro-inflammatory cytokines and inhibit the effects of T helper cells, thus preventing further inflammation, oxidative stress and placental dysfuntion^69^. *PZP* is also highly expressed in late-pregnancy serum and its upregulation during pregnancy represents a major maternal adaptation that helps to maintain extracellular proteostasis during gestation^68^. Furthermore, balanced expression of Glycodelin A (GdA) and its carrier protein *PZP* in the decidua seems crucial for a successful ongoing pregnancy^70^. Even though we cannot pinpoint putative causal variants, our study provides important additional support for the pathophysiological role of *PZP* in preeclampsia.

The *ACTN4* gene encodes for an actin-binding protein with multiple roles in different cell types, and is well known for causing focal segmental glomerulosclerosis^71–73^. The gene product is also involved in the maintenance of cytoskeletal structure, modulating cell motility and regulating endothelial cells^74,75^, and may play a role in placentation^76^. *ACTN4* has been suspected to act as a regulator of trophoblast proliferation and differentiation during early pregnancy, and has been shown to have reduced expression in severe preeclampsia^76^. Specifically, the lack of expression appears to prevent cytotropboblast differentiation into interstitial extravillous cytotrophoblasts, possibly compromising the remodelling of the spiral arteries. In addition, the reduced *ACTN4* levels have been suggested to activate endothelial cell apoptosis^77^. In laboratory studies conducted by Kos et al.^73^, *ACTN4*^-/-^ mice developed a severe glomerular disease with no other histological abnormalities. However, only 6 % of the offspring of mated *ACTN4*^+/-^ mice were homozygous for the dysfunctional gene, instead of the expected 25 %. Whether the reduced litter size observed in this study was caused by dysfunctional placental development remains unclear since the placentas were unfortunately not examined. Further functional studies are needed to gain evidence of the potential causal role of *ACTN4* in preeclampsia and to uncover pathophysiological mechanisms behind the association of this locus.

PE-HTP-META also replicated four out of the five associations identified in Steinthorsdottir et al.^11^, with only the locus in 12q24 remaining slightly below genome-wide significance.

### PE-FGR-META reveals putative association with uterine natural killer cells

Two loci were associated with the PE-FGR phenotype with genome-wide significance. The first replicated the association near *FGF5* (rs16998073) and the second overlapped the novel locus near the *AUH* and *LINC00484* genes that was also detected in PE-META, but remained without a plausible biological explanation. Interestingly, the lead variant of the latter finding in PE-FGR-META is rs7470773, which is in only modest LD (r^2^ 0.34) with the PE-META lead variant rs7862828, suggesting that the mechanisms behind the association might differ between the two phenotypes. Functional characterization of the locus revealed an interesting intergenic variant rs7028982 (p-value 6.7e-09, beta 0.13) that is in tight LD (r2 0.98) with the lead variant rs7470773. This variant is positioned within a highly conserved genomic element (phastCons score 417). The allele associated with increased PE-FGR risk reduces the expression of two nearby poorly characterized long non-coding RNA genes *LINC00484* and ENSG00000273381 in several tissues, but the allele also reduces the expression of *NFIL3* in whole blood (normalized effect size -0.14). In mice studies, the dysfunction of *NFIL3* on 9q22 has been shown to cause absence of uterine natural killer cells^78^, which have a central role in the placental vascular remodeling during pregnancy^79^. This absence results in incomplete remodeling of the uterine arteries and decidua, placental defects, and fetal growth restriction^78^, which are all features associated with preeclampsia.

### Paternal and fetal association analyses in the FINNPEC cohort

GWAS of paternal and fetal PE, PE-HTP and PE-FGR did not yield any genome-wide significant associations (Figure S6). Uncovering the paternal and fetal associations may require significantly larger sample sizes than available in the current study.

## Discussion

In this largest GWAS meta-analysis of maternal preeclampsia to date, we identified multiple novel risk loci for the strict preclampsia and for the broader phenotypes of hypertensive pregnancy. By combining the GWAS summary statistics from the Finnish and Estonian study populations we received a large and ethnically relatively homogenous data set. For the strict preeclampsia phenotype, we supplemented the Finnish-Estonian data with the summary statistics from the large and genetically more diverse InterPregGen study (Steinthorsdottir et al.) to increase the statistical power. In preclampsia, general hypertensive pregnancy and preeclampsia-SGA phenotypes we identified altogether 18 genome-wide significant loci, 12 of which had not previously been reported in the genome-wide studies of maternal preeclampsia. These results offer valuable insights into the genetic architecture and biology behind preeclampsia as well as into the connection between preeclampsia and related pregnancy phenotypes such as gestational hypertension and impaired fetal growth.

### Novel associations shed light on the genetic architecture of preeclampsia

As the life of every new individual begins with fetal development and birth, it is an important period also from an evolutionary perspective. All dysfunctions in this process have the potential of severely reducing the biological fitness (i.e. the likelihood of future reproductive success of the individual) of the mother and the offspring. The evolutionary origin of preeclampsia has been suggested to be related to the particularly large brain size of humans, which imposes strict requirements for efficient oxygen flow during pregnancy, and thus, deep implantation of the placenta to the uterus^80^. However, this places great stress on the maternal circulation. Such opposing selection patterns is called antagonistic pleiotropy – a phenomenon which has been shown to be enriched in the coronary artery disease related genes^81^. Specific HLA-G haplotypes were lately suggested as being under balancing selection, with heterozygotes providing resistance towards infectious diseases, but male fetal homozygotes resulting in maternal immune response and increased preeclampsia risk^38^. Interestingly, a recent study^53^ found several genes that are associated with preeclampsia in our study – namely *FGF5, SH2B3, FTO* and the HLA region – to be amongst the reported 20 most pleiotropic. Likewise, the other genes that probably drive the associations found in our study appear to affect several different traits, as discussed in the results and more in depth in the following paragraphs.

Theoretically, in pleiotropic scenarios, the loci of small effects are expected to have pronounced contribution, as the loci of large effect have a deleterious impact, and thus become under negative selection, eventually purging them from the population^82^. Absence of large effect susceptibility variants has been detected in early onset diseases, such as type I diabetes, but not for instance in cancers^83^. This purging of high effect alleles from the population likely inflates the polygenic nature of the disorder^84^. Such genetic architecture complicates pinpointing the causal variants, as detecting weakly associated alleles in GWAS requires large samples sizes^85^. Indeed, earlier genome-wide association studies on preeclampsia have yielded few findings, despite including fairly large number of samples. The ongoing era of GWAS has permitted generating novel hypotheses on molecular background of numerous medical conditions over the past decade^86^ – but preeclampsia related findings have been few and far between. The results of our meta-analysis demonstrate that we now begin to reach large enough sample size to finally utilize the benefits of GWAS in the study of preeclampsia to uncover its underlying genetic architecture.

### Findings underline the prominent role of factors related to cardiometabolic, endothelial and placental dysfunction

The 6/9 loci in the PE phenotype, 11/13 HTP loci and 1/2 PE-FGR loci have reached genome-wide significant association in the previous GWA studies of cardiovascular diseases. This strongly implies that the established blood pressure loci modify predisposition to hypertension also during pregnancy, plausibly via the same mechanisms. This interpretation is also supported by our genetic correlation results: correlations of preeclampsia with blood pressure medication and high blood pressure were as high as 0.587 and 0.560, respectively. Moreover, correlations between preeclampsia and stroke, heart attack and coronary artery disease were all above 0.4. These results are in accordance with the InterPregGen study which reported that the polygenic risk score of hypertension was associated with preeclampsia. In addition, the well-established epidemiological evidence of the shared risk factors between preeclampsia and cardiovascular diseases as well as the increased incidence of cardiovascular diseases after preeclampsia further support the existence of the shared genetic risk factors for these conditions^13–15^. All in all, the findings of our study support the idea of pregnancy as a window to future cardiovascular health: the increased genetic susceptibility to cardiovascular disease might become evident for the first time during pregnancy.

While this study does not reveal the putative causal variants in many of the loci, we are able to recognize general trends when looking at most likely candidate genes in each locus. We are now able to show genome-wide significant associations between the hypertensive pregnancy disorders and the loci related to endothelial dysfunction, placental development and immunology. Systemic endothelial dysfunction is central to the pathophysiology of preeclampsia and is characterized by impaired vasodilation, endothelial injury, and reduction in vascular integrity^87–89^. The best known biomarker of preeclampsia, sFlt-1, is a soluble antiangiogenic protein that reduces the availability of the proangiogenic proteins *VEGF* and *PlGF* to endothelial cells therefore impairing the maintenance of vascular integrity and cellular viability^90,91^. Our study provides strong hypothesis-free evidence of the relevance of the *FLT1* gene in maternal preeclampsia. Interestingly, also several other genes proximal to the associated lead SNPs of the current study, including *NPPA* on 1p36, *FES* and *FURIN* on 15q26, *ACTN4* on 19q13, and and *PREX1* on 20q13.13, encode for proteins that are involved in regulating endothelial permeability and leukocyte transmigration^92–96^. These findings provide further support to the idea that preeclampsia liability might be modified by alterations in the integrity of the endothelium.

Abnormal leakage of protein to urine is another key feature of preeclampsia. Intriguingly, several of the genes proximal to our associating lead SNPs have been linked with kidney disease. Mutations in *PLCE1, TNS2, ACTN4*, and *TRPC6* cause nephrotic syndrome characterized by proteinuria^57,72,97–100^. Products of these genes have important roles in podocyte function and integrity of the glomerular filtration barrier^71,101–103^. The mechanisms of action of these genes in causing kidney damage are likely to be variable. For example, *TRPC6* might contribute to the podocyte damage in preeclampsia by mediating the effects of angiotensin II type I receptor agonistic autoantibody^104^, which is also known to increase the effect of angiotensin II leading to vasoconstriction and activation of reactive oxygen species.

Many of the genes next to the lead SNPs of our study are involved in the placental development and function, which are often compromised in preeclampsia. Both of the genes closest to the lead variant in the locus 11q22, *PGR* and *TRPC6*, as well as *NFIL3* on 9q22, are known to affect placental functions and maintenance of pregnancy, as described earlier. *PGR* has been suggested to contribute to balanced hormonal signalling during pregnancy, and subsequently aid the immune and endothelial cells in the cytotrophoblast invasion^56^. Also the 19q13 located *ACTN4* is known for regulating the trophoblast proliferation and differentiation during early pregnancy. Specifically, the lack of *ACTN4* expression appears to prevent the cytotropboblast differentiation into the interstitial extravillous cytotrophoblasts, possibly compromising the remodelling of the spiral arteries^76^. During placentation, the invasive trophoblast cells called the extravillous trophoblasts migrate into the maternal spiral arteries and, in cooperation with the maternal cells, transform these arteries into dilated and non-contractile tubes capable of providing adequate perfusion for the growing fetus. Defects in these processes are well-documented especially in the early-onset preeclampsia^3,4^.

Immunological factors have been shown to contribute to pathophysiology of preeclampsia. In this work, the locus on HLA region is particularly intriguing. Even though this work is not able to pinpoint the causal variant or gene, the association is possibly driven by variants in the psoriasis causing candidate genes *CCHCR1* or *PSORS1C2*. Additional examples of genes putatively modulating the immunological response to pregnancy include *NFIL3* (a candidate gene in the PE and PE-FGR phenotypes), suspected to contribute to the numbers of uterine natural killer cells, and *PZP* (in the PE-HTP phenotype) likely modulating T helper cell response. Fetal growth restriction is an indication of severe preeclampsia^105^, and immunological etiology is typically linked to patients with severe/early-onset preeclampsia^106^. Immunology related findings have been absent in previous preeclampsia GWAS results, but our work provides the first support for their role in this type of studies.

Examples of pleiotropic candidate genes of the associating loci in our study include the natriuretic peptide genes *NPPA* and *NPPB* on 1p36 and their clearance receptor *NPR3* on 5p13. Natriuretic peptide hormones regulate blood pressure and renal function among their several other effects^107^. In addition, mice that lack the expression of atrial natriuretic peptide develop gestational hypertension and proteinuria and, similar to preeclampsia, exhibit impairment in trophoblast invasion and uterine spiral artery remodeling^108,109^. It is tempting to hypothesize that changes in the function or expression level of these genes with several effects in the key mechanisms of preeclampsia could contribute to the multi-organ dysfunction that is characteristic of this pregnancy disorder. As we were not, however, able to pinpoint the actual causal variants in the associating loci, further research is required to verify the relevance of the candidate genes highlighted by our study.

The exploration of combinatory phenotypes that included also other hypertensive disorders of pregnancy and, on the other hand, fetal growth restriction in addition to preeclampsia, abled us to estimate the connections between these related disorders. The effect sizes of the uncovered loci were largely similar between the different phenotypes explored in this study. However, when comparing genome-wide significant loci between the PE, PE-HTP and PE-FGR phenotypes, the most likely candidate genes on many PE-HTP loci appeared to be related to hypertensive diseases. This observation implies that preeclampsia, gestational hypertension and preeclampsia superimposed on chronic hypertension share genetic risk factors with each other, and the same factors also increase the risk of hypertension later in life. Some of the previously identified hypertension risk loci were also associated with the PE and PE-FGR phenotypes, but in these two phenotypes the number of the associations with the known hypertension loci was smaller and the results also pointed towards other processes, such as the placental development and immunology. Future studies with even larger sample sizes and detailed phenotype information will aid in classifying pregnancy disorders in general and subtypes of preeclampsia in specific.

The main strength of this study is the use of three well characterized cohorts of FINNPEC, FinnGen and EstBB, originating from two closely related populations of Finns and Estonians. We also perform the largest genetic study of preeclampsia to date, when we include the results from the previous InterPregGen study. By comparing the PE-HTP and PE-FGR phenotypes we are able to obtain a more nuanced view on possible genetic variants contributing to preeclampsia phenotypes leaning towards either hypertensive disorders of pregnancy or towards placental defects. The register-based approach used in this study can be seen as a limiting factor, as FinnGen and the EstBB do not contain identifying information of the subjects and thus we are not able to directly validate the ICD-codes. Nonetheless, the coverage and accuracy of the Finnish Care Register for Health Care has previously been shown to be excellent^110^. Furthermore, our PE and PE-HTP phenotypes replicate the findings of the largest previously published GWAS meta-analysis and our different study cohorts provide uniform support for our findings with no evidence of heterogeneity.

In conclusion, our study uncovers 12 novel loci with genome-wide significant association with preeclampsia or hypertensive disorders of pregnancy. We show that the cardiovascular disease related genes have a central role in the pathophysiology of preeclampsia as previously suggested, but importantly we also emphasize that many of those genes have pleiotropic effects on cardiometabolic, endothelial and placental function. In addition, we provide further evidence for a role of several loci not previously associated with cardiovascular disease, but containing genes with apparent importance in the maintenance of successful pregnancy, with dysfunctions leading to preeclampsia-like symptoms.

## Supporting information

Suppelementary

## Data Availability

Full meta-analysis summary statistics will be made available upon publication.

## Acknowledgements

We appreciate contribution of the members and assisting personnel of the FINNPEC study. The FINNPEC study was supported by the Jane and Aatos Erkko Foundation, Juho Vainio Foundation, Päivikki and Sakari Sohlberg Foundation, Academy of Finland, research funds of the University of Helsinki, government special state subsidy for the health sciences for the Hospital District of Helsinki and Uusimaa, Finska Läkaresellskapet, Liv och Hälsa Foundation, NovoNordisk Foundation, Finnish Foundation for Pediatric Research, Emil Aaltonen Foundation, Sigrid Juselius Foundation, Signe and Ane Gyllenberg Foundation and Finnish Foundation for Laboratory Medicine. We wish to thank InterPregGen consortium members for assistance in accessing the summary data of Steinthorsdottir et al. study^11^. We appreciate the contribution of the EstBB participants and staff.

We want to acknowledge the participants and investigators of FinnGen study. The FinnGen project is funded by two grants from Business Finland (HUS 4685/31/2016 and UH 4386/31/2016) and the following industry partners: AbbVie Inc., AstraZeneca UK Ltd, Biogen MA Inc., Bristol Myers Squibb (and Celgene Corporation & Celgene International II Sàrl), Genentech Inc., Merck Sharp & Dohme Corp, Pfizer Inc., GlaxoSmithKline Intellectual Property Development Ltd., Sanofi US Services Inc., Maze Therapeutics Inc., Janssen Biotech Inc, Novartis AG, and Boehringer Ingelheim. Following biobanks are acknowledged for delivering biobank samples to FinnGen: Auria Biobank (www.auria.fi/biopankki), THL Biobank (www.thl.fi/biobank), Helsinki Biobank (www.helsinginbiopankki.fi), Biobank Borealis of Northern Finland (https://www.ppshp.fi/Tutkimus-ja-opetus/Biopankki/Pages/Biobank-Borealis-briefly-in-English.aspx), Finnish Clinical Biobank Tampere (www.tays.fi/en-US/Research_and_development/Finnish_Clinical_Biobank_Tampere), Biobank of Eastern Finland (www.ita-suomenbiopankki.fi/en), Central Finland Biobank (www.ksshp.fi/fi-FI/Potilaalle/Biopankki), Finnish Red Cross Blood Service Biobank (www.veripalvelu.fi/verenluovutus/biopankkitoiminta) and Terveystalo Biobank (www.terveystalo.com/fi/Yritystietoa/Terveystalo-Biopankki/Biopankki/). All Finnish Biobanks are members of BBMRI.fi infrastructure (www.bbmri.fi). Finnish Biobank Cooperative -FINBB (https://finbb.fi/) is the coordinator of BBMRI-ERIC operations in Finland. The Finnish biobank data can be accessed through the Fingenious^®^ services (https://site.fingenious.fi/en/) managed by FINBB.

The Genotype-Tissue Expression (GTEx) Project was supported by the Common Fund of the Office of the Director of the National Institutes of Health, and by NCI, NHGRI, NHLBI, NIDA, NIMH, and NINDS. The data used for the analyses described in this manuscript were obtained from the GTEx Portal on 03/24/22.

