## Supplementary material for "GWAS of preeclampsia and hypertensive disorders of pregnancy uncovers genes related to cardiometabolic, endothelial and placental function": Suppelementary

**Supplementary Table S1**. ICD codes for the FinnGen phenotypes of hypertensive pregnancy.

| Phenotype | ICD-10 | ICD-9 | ICD-8 |
|---|---|---|---|
| PE | O11, O14.0, O14.1, O14.9, O15.0, O15.1, O15.2 | 6424, 6425, 6426, 6427 | 63703, 63704, 63709, 63710, 63799, 66120 |
| HTP | O10, O11, O13, O14, 015, O16 | 642 | 63701, 63703, 63704, 63709, 63710, 63799, 66120 |
| PE-SGA | O11, O14.0, O14.1, O14.9, O15.0, O15.1, O15.2, O36.5 | 6424, 6425, 6426, 6427, 6565A, 6565B | 63703, 63704, 63709, 63710, 63799, 66120 |

HTP=hypertensive pregnancy; PE=pre-eclampsia, PE-SGA=pre-eclampsia or small-for gestational age baby. Control groups for each of the phenotypes consisted of parous women with no case ICD codes for the phenotype in question.

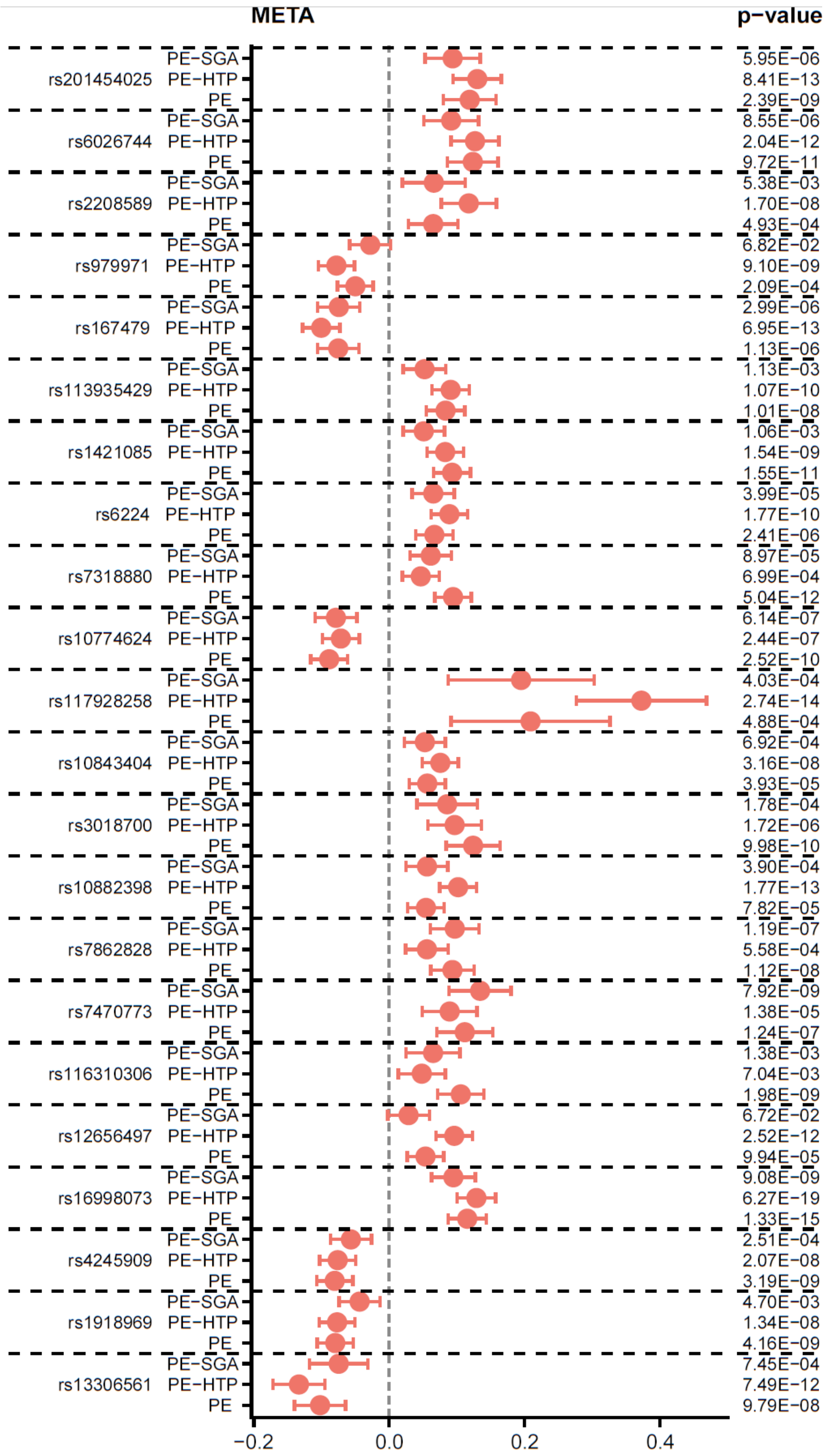


Figure S1. Forest plot of meta-analysis results for each different phenotype. PE=preeclampsia; PE-HTP=preeclampsia, gestational hypertension or preeclampsia superimposed on chronic hypertension; PE-SGA=preeclampsia or indication of fetal growth restriction.

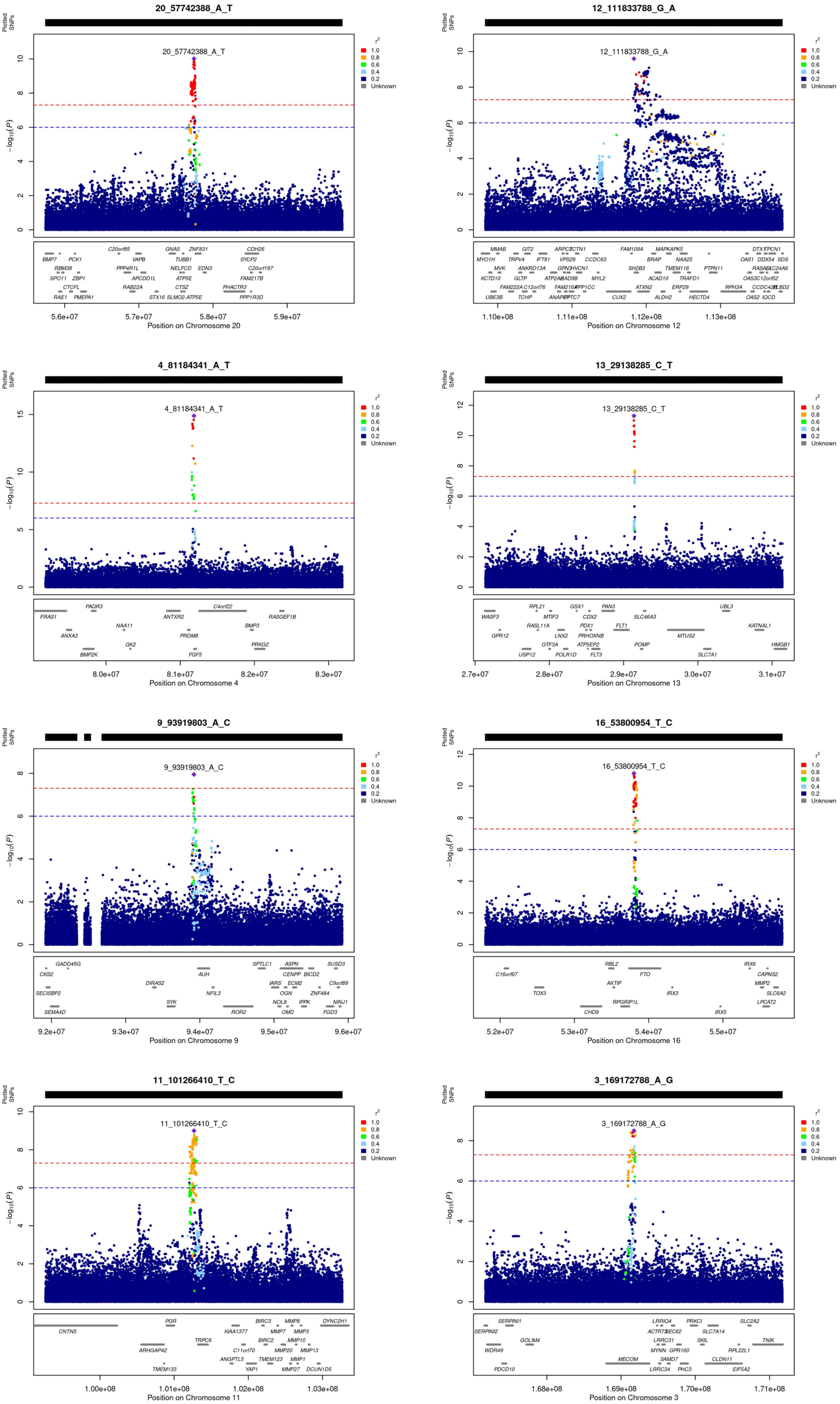


*Figure S2. Regional association plots for genome wide significant associations of PE-META. HLA-region figure could not be produced by the 'locuszooms' software due to high gene density.*

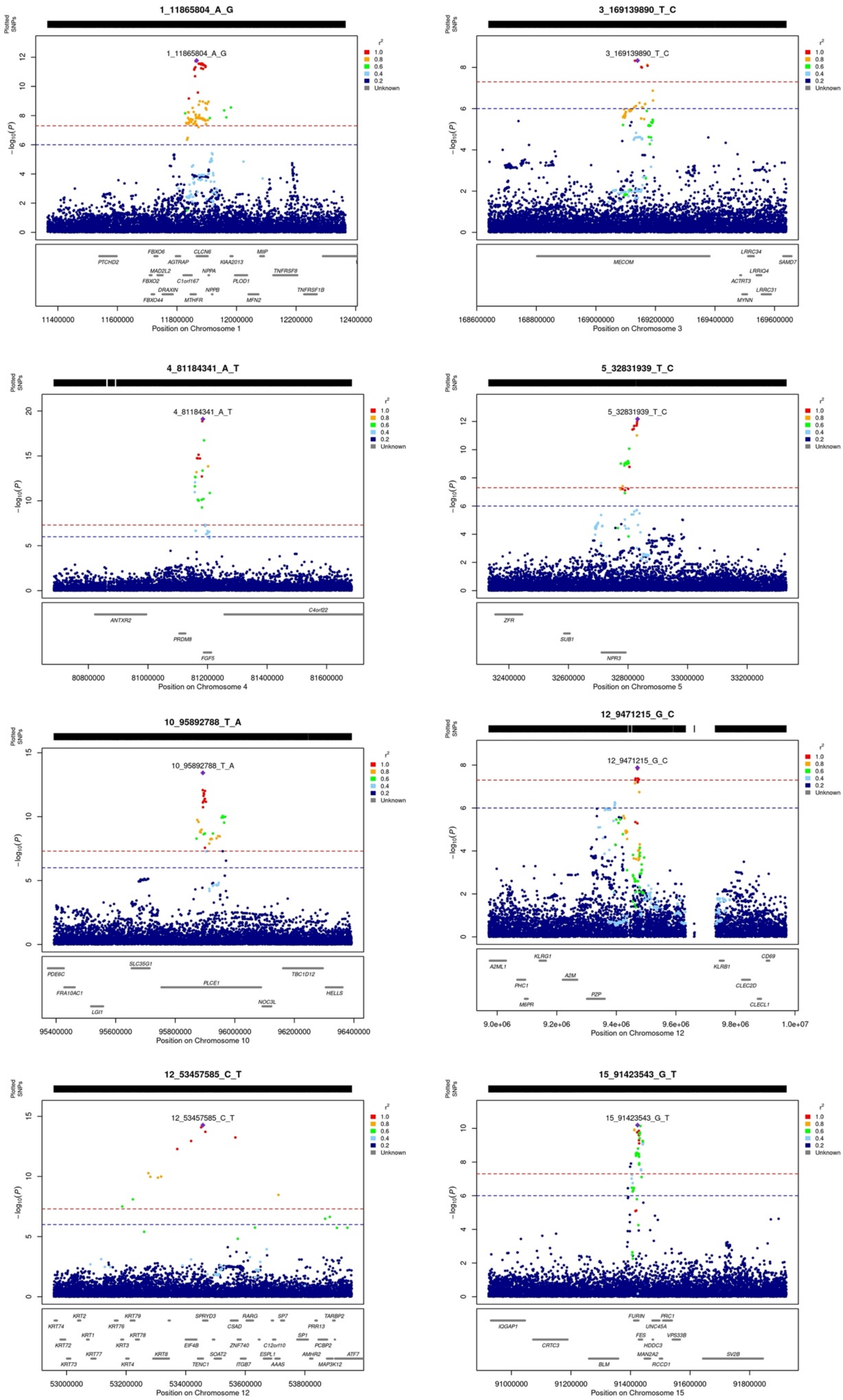


*Figure S3. Regional association plots for genome wide significant associations of PE-HTP-META for chromosomes 1-15. PE-HTP=preeclampsia, gestational hypertension or preeclampsia superimposed on chronic hypertension.*

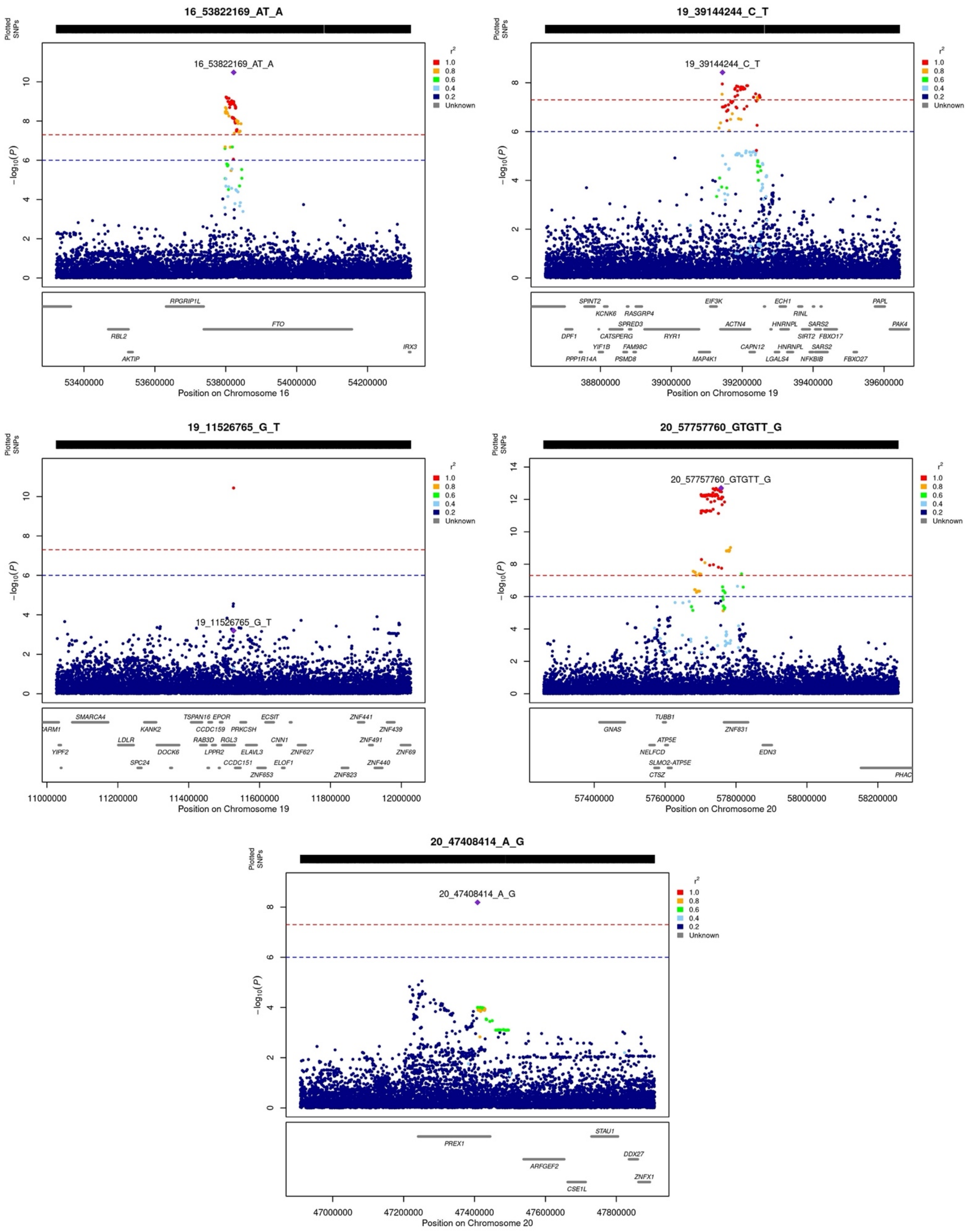


*Figure S4. Regional association plots for genome wide significant associations of PE-HTP-META for chromosomes 16-20. PE-HTP=preeclampsia, gestational hypertension or preeclampsia superimposed on chronic hypertension.*

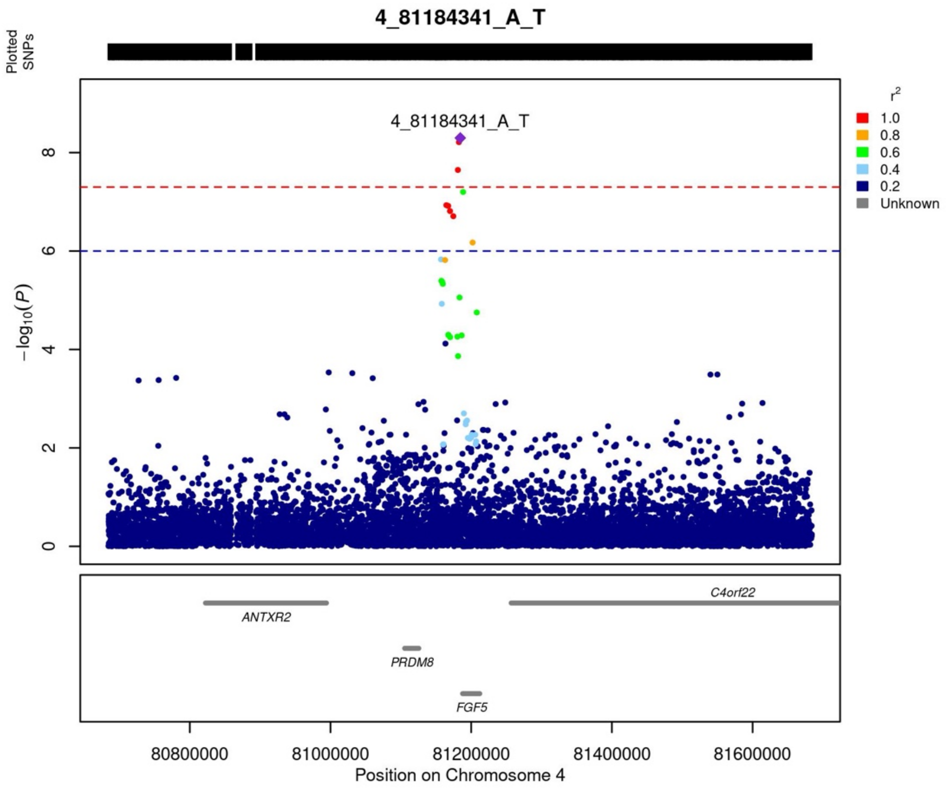


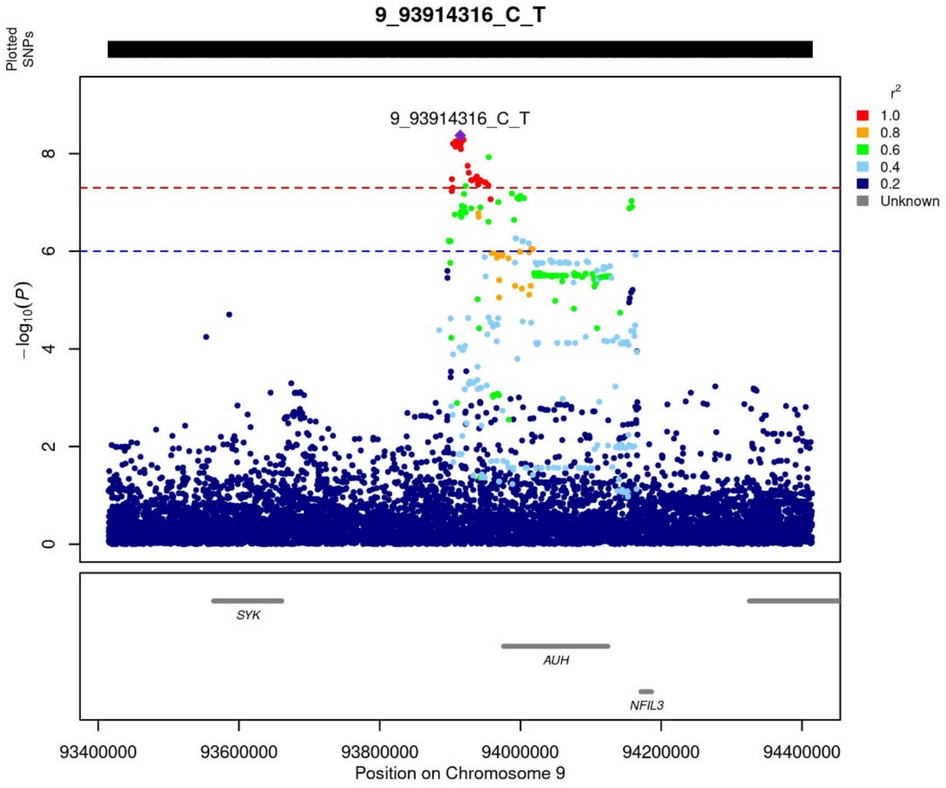


*Figure S5. Regional association plot for PE-SGA lead variants. PE-SGA=preeclampsia or indication of fetal growth restriction.*

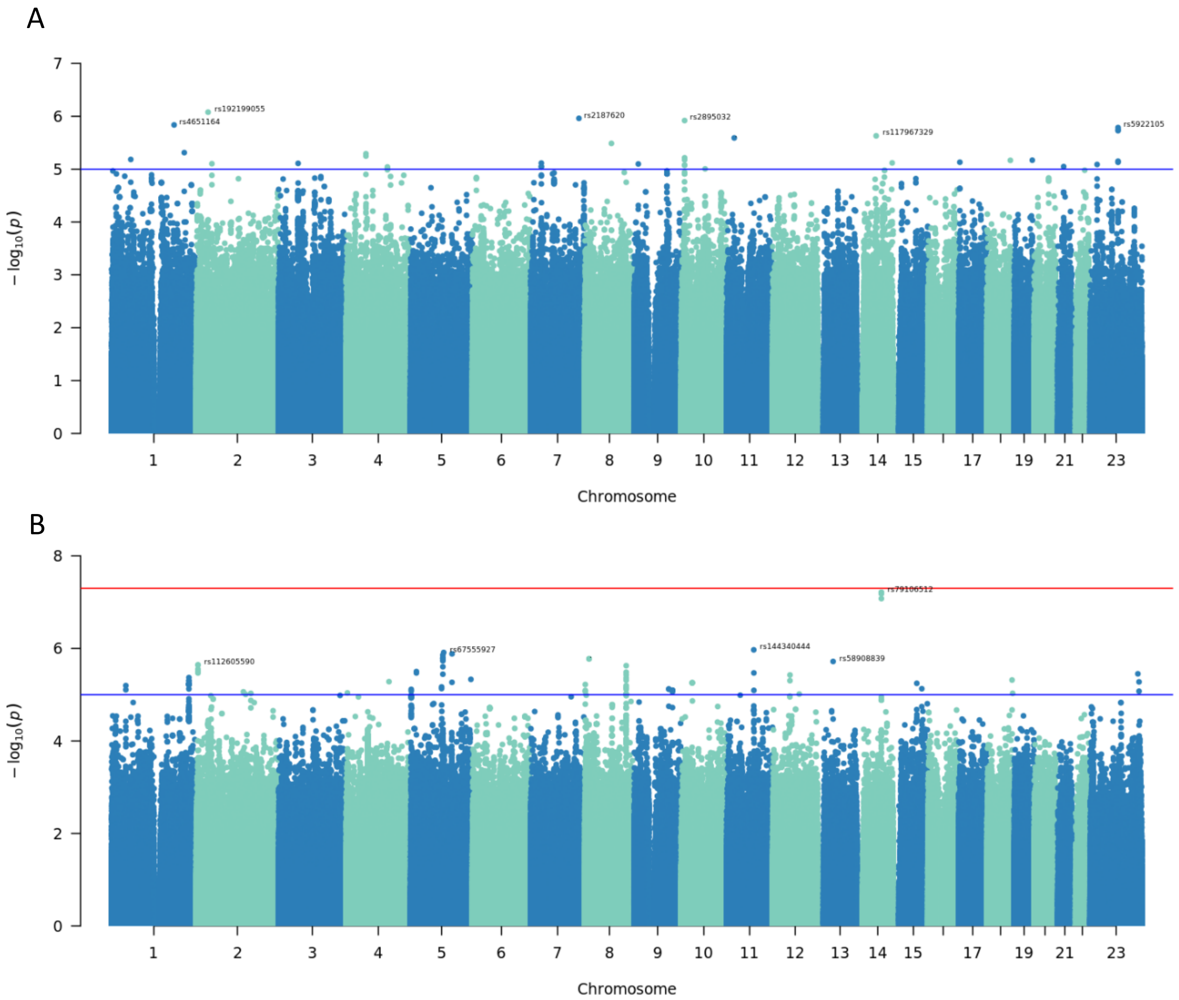


*Figure S6. Regional association plot for PE lead variants in paternal (A) and child (B) samples.*
